# Mental-state reasoning or downstream vascular burden? Theory of Mind task performance in post-stroke aphasia

**DOI:** 10.64898/2026.04.14.26350532

**Authors:** J. Kurtz, A. Billot, I. Falconer, H. Small, A. Charidimou, S. Kiran, M. Varkanitsa

## Abstract

**Background:** Theory of Mind (ToM) deficits are well-documented in right-hemisphere stroke but remain understudied in post-stroke aphasia. Prior work suggests that performance on tasks assessing ToM may be relatively preserved in aphasia and dissociable from language impairment, but these findings are based largely on small studies. This study examined performance on nonverbal false-belief tasks in post-stroke aphasia, its relationship with aphasia severity, and whether vascular brain health, operationalized using cerebral small vessel disease (CSVD) markers, contributed to variability in performance.

**Methods:** Forty-four individuals with aphasia completed two nonverbal belief-reasoning tasks assessing spontaneous perspective-taking and self-perspective inhibition. Task accuracy served as the primary outcome. Linear regression models examined associations between task performance, aphasia severity (Western Aphasia Battery-Revised Aphasia Quotient), and CSVD markers, including white matter hyperintensities, cerebral microbleeds, lacunes and enlarged perivascular spaces in the basal ganglia and centrum semiovale.

**Results:** Performance was heterogeneous across tasks, with reduced performance observed in 23% of participants on the Reality-Unknown task and 36% on the Reality-Known task. Aphasia severity was not associated with task accuracy. Greater cerebral microbleed count was associated with lower accuracy on both tasks, while greater basal ganglia enlarged perivascular spaces burden showed a more selective association with lower performance.

**Conclusions:** Performance on nonverbal false-belief tasks in aphasia is variable and not explained by aphasia severity alone. These findings suggest that apparent ToM-related difficulties in aphasia may be shaped by broader vascular brain health, supporting a more multidimensional framework for interpreting social-cognitive task performance after stroke.

## Introduction

Theory of Mind (ToM)—the ability to attribute mental states such as beliefs, desires, and intentions to others—is a key aspect of social cognition that allows individuals to predict and interpret behavior. Early lesion studies and reviews linked ToM impairments primarily to right-hemisphere damage,^1–3^ whereas individuals with left-hemisphere lesions often perform comparably to neurotypical controls. This pattern contributed to the longstanding assumption that ToM is largely right lateralized. More recent functional neuroimaging work, however, suggests a broader and more distributed mentalizing network, centered on the right temporoparietal junction but also involving medial prefrontal and posterior midline regions, and in many paradigms bilateral temporoparietal cortex^4^. Social cognition has often remained peripheral to the study of aphasia, even though individuals with aphasia must navigate social interactions while living with both neurological injury and language impairment.

The small literature available in aphasia suggested that individuals with aphasia can succeed on classic ToM tasks such as false-belief reasoning, that is, understanding that another person can hold a belief that differs from reality and from one’s own knowledge.^5, 6^ Two influential single-case studies reported preserved performance on first-order false-belief tasks in older adlts with left-hemisphere damage and severe language impairment.^7, 8^ Crucially, both individuals performed well on picture-based tasks designed to minimize linguistic demands. These findings have often been interpreted as evidence that social-cognitive reasoning can remain preserved in aphasia, suggesting at least partial dissociation between the neural systems supporting language and those supporting mental-state attribution. At the same time, these studies were based on single-cases and may not capture the full spectrum of performance observed across individuals with aphasia, particularly given the heterogeneity of aphasia profiles and the multiple cognitive operations required for successful task completion.^9, 10^

Understanding how individuals with aphasia perform on tasks intended to index mental state reasoning is important for refining brain–behavior models of communication, as well as for understanding functional communication, social participation, and overall quality of life.^11^ Difficulties on such tasks may be clinically meaningful even if they do not necessarily reflect a primary impairment in ToM itself, given that performance can be shaped by executive demands as we will discuss in the following paragraph. Everyday social interaction depends on the ability to track perspectives, interpret communicative intent, and respond flexibly to others’ behavior. When these processes are disrupted, conversation flow may break down, relationships may become strained, and misunderstandings or social withdrawal may follow.^12, 13^ These difficulties may go unrecognized when clinical attention is focused primarily on overt language production and comprehension, rather than on the broader social-cognitive demands of interaction. As a result, individuals with aphasia may experience reduced opportunities for meaningful interaction, even when their verbal output appears relatively preserved.^11^

In this study, we examine performance on two nonverbal false belief tasks^14^ in a larger group of individuals with aphasia. Although these tasks place minimal demands on language, successful performance may still depend on executive processes such as inhibition, working memory, and cognitive flexibility, which support the ability to maintain, update, and suppress competing perspectives. Prior work suggests that executive functions contribute importantly to performance on many ToM tasks, although the extent to which these processes are intrinsic to ToM itself, rather than support processes required by particularly tasks, remains debated^15–17^. This distinction is particularly relevant in aphasia. Even when linguistic demands are reduced, performance on ToM tasks may still be shaped by disruption of broader domain-general cognitive systems. Accordingly, individual differences on these tasks may reflect not only the effects of focal left-hemisphere stroke, but also the integrity of more distributed neural systems that support executive domain-general cognition. This makes overall brain health, broadly defined as the structural and functional integrity of neural systems that support cognition, emotion, and behavior,^18^ highly relevant for interpreting apparent ToM-related difficulties in aphasia.

Brain health can be influenced by several factors, including aging and vascular pathology, both of which may affect the executive processes that support performance on ToM tasks. Aging, in particular, is associated with gradual declines in executive functions such as inhibition, cognitive flexibility, and working memory, which in turn have been linked to poorer performance on cognitively demanding ToM tasks, especially those requiring inhibitory control and perspective shifting.^19–22^ Less is known about how vascular pathology might impact ToM abilities. A common form of vascular pathology in the elderly is cerebral small vessel disease (CSVD), a degenerative microvascular process that leads to a range of structural and functional brain alterations detectable on neuroimaging.^23^ Importantly, CSVD is highly prevalent in stroke survivors, including those with aphasia following left hemisphere stroke, and is the major contributor to vascular cognitive impairment and dementia. ^24, 25^ In addition to its well-established effects on attention, executive function, and memory, emerging evidence suggests that CSVD may also compromise social-cognitive domains. For instance, extensive white matter hyperintensities, a hallmark of CSVD, have been linked to lower ToM task performance in community-dwelling older adults.^26^ These findings raise the possibility that vascular burden, beyond focal stroke-related damage, may contribute to variability in ToM performance in individuals with aphasia.

The present study therefore examines performance on nonverbal false-belief tasks in individuals with aphasia with three aims: (1) to characterize the frequency and distribution of low performance on these tasks in this population, (2) to determine whether task performance varies as a function of aphasia severity, and (3) to investigate whether CSVD contributes to variability in task performance beyond focal stroke lesion burden and aphasia severity. We predicted that reduced performance would be observed in at least a subset of individuals with aphasia. Because performance on these tasks may also be shaped by executive demands, such findings would require cautious interpretation and would not, on their own, establish a primary ToM deficit. We further predicted that aphasia severity alone would not fully account for variability in performance, given the reduced linguistic demands of the tasks. Finally, we predicted that greater CSVD burden would be associated with poorer performance, particularly under conditions that place greater demands on executive control. Together, these aims support a broader framework for understanding apparent ToM-related difficulties in aphasia—one that interprets them not simply as a consequence of language impairment or as direct evidence of impaired mental-state reasoning, but as a downstream behavioral manifestation of overall brain health, including vascular burden.

## Methods

### Participants

Forty-four individuals with chronic post-stroke aphasia due to left hemisphere stroke participated in this study. Participants were fluent English speakers, had normal or corrected to normal hearing and vision, were medically and neurologically stable, and had received at least a high school education. Summary demographic and clinical characteristics are provided in Table 1, and individual-level participant data are reported in Supplementary Table 1. All participants provided written informed consent in accordance with the Boston University Institutional Review Board and the Massachusetts Institute of Technology’s Committee on the Use of Humans as Experimental Subjects.

**Table 1.** Participant Characteristics.

| Variable | Mean (SD) | Range | N |
| --- | --- | --- | --- |
| Age (years) | 59.07 (10.57) | 20 – 83 | 44 |
| WAB-R AQ | 81.84 (18.20) | 31.5 – 100 | 44 |
| Years of Education | 15.84 (2.61) | 12 – 22 | 44 |
| Lesion Volume (mm<sup>3</sup>) | 88,447.83 (74,943.04) | 2,025 – 333,891 | 39* |
| MPO (months) | 80.75 (83.47) | 5 – 539 | 44 |
| Gender | 32 Male (73%), 12 Female (27%) | — | 44 |
| Race | 38 White (86%), 6 Black/African American (14%) | — | 44 |
| Ethnicity | 41 Non-Hispanic (93%), 3 Hispanic (7%) | — | 44 |

WAB-R AQ: Western Aphasia Battery-Revised Aphasia Quotient; MPO: Months Post-Onset
\* Fewer participants had lesion volume data available due to missing MRI scans

**Table 2.** Cerebral Small Vessel Disease Markers in the Study Cohort.

| **CSVD Marker** | **Summary Statistic** | **Range** | **N** |
| --- | --- | --- | --- |
| Periventricular WMH (Fazekas 0–3) | Median = 1 (IQR: 1–2) | 0–3 | 31 |
| Deep WMH (Fazekas 0–3) | Median = 1 (IQR: 0–2) | 0–3 | 31 |
| Basal Ganglia EPVS (0–4) | Median = 2 (IQR: 1–2) | 0–4 | 31 |
| Centrum Semiovale EPVS (0–4) | Median = 2 (IQR: 2–3) | 1–4 | 31 |
| Lacunes (presence) | 8 present (24%) | — | 31 |
| Lacunes count | Median = 0 (IQR: 0–1) | 0–3 | 31 |
| CMB (presence) | 9 present (27%) | — | 32 |
| CMB count | Median = 0 (IQR: 0–1) | 0–7 | 32 |
| Total CSVD score (0–4) | Median = 1 (IQR: 1–2) | 0–3 | 28 |

WMH: white matter hyperintensities; EPVS: enlarged perivascular spaces; CMB: cerebral microbleeds; CSVD: cerebral small vessel disease; IQR = interquartile range (25th–75th percentile)

### Behavioral Data

Aphasia severity was assessed with the Western Aphasia Battery-Revised Aphasia Quotient (WAB-R AQ).^27^ ToM was assessed using two nonverbal false-belief reasoning tasks designed to minimize language demands while isolating belief reasoning.^14^ The *Reality-Unknown* (RUK) task assessed spontaneous false-belief reasoning under low inhibitory demands. Participants viewed short, silent videos in which a character observed the location of a hidden object while the participant did not. In false-belief trials, the character left the scene and the boxes containing the object were swapped in her absence. Upon returning, the character placed a marker on the box corresponding to her outdated belief. Participants were asked to indicate the actual location of the object. Correct performance required inferring the character’s false belief and using this inference to deduce the object’s true location. Crucially, because participants never directly observed the object’s initial placement, belief inference occurred in the absence of competing self-knowledge, minimizing inhibitory demands. The *Reality-Known* (RK) task assessed false-belief reasoning under high inhibitory demands. Participants viewed short, silent videos in which both they and the character observed the initial placement of an object. In false-belief trials, the character exited the scene, and the object was visibly moved to a new location. Upon return, the character remained unaware of the switch. Participants were asked to indicate where the character would look for the object. Correct responses required participants to inhibit their own knowledge of the actual location and reason about the character’s mistaken belief. Both tasks included control trials (true belief, memory control, and filler) to assess non-social reasoning and discourage superficial strategies. We used the decision trees developed in the original study to classify responses to impaired versus spared belief reasoning based on performance across conditions and normative patterns.^14^ We also calculated overall percent accuracy across all trials.

### Neuroimaging and CSVD Scoring

For a subset of participants (n = 32) multi-sequence imaging (T1 MPRAGE, T2-SPACE FLAIR, SWI) was available to evaluate CSVD markers, including white matter hyperintensities (WMH), enlarged perivascular spaces (EPVS), lacunes, and cerebral microbleeds (CMB). Rating was conducted by two raters blinded to all patients’ characteristics according to the standards for reporting vascular changes on neuroimaging consensus criteria^28^. A total CSVD score was also calculated. Full imaging parameters, rating criteria, and scoring procedures are detailed in the Supplement.

### Statistical Analysis

To examine the relationship between aphasia severity and nonverbal ToM performance, linear regression models were used with overall percent accuracy across conditions of each ToM task as the dependent variable. Separate models were run for the RUK and RK tasks, with aphasia severity (WAB-R AQ) as the primary predictor. These models were also adjusted for age and lesion volume. To assess the role of brain health, separate linear regression models tested associations between ToM percent accuracy and individual CSVD markers, including PVWMH and DWMH, EPVS, lacunes, CMB, and a total CSVD burden score, while controlling for age and lesion volume. Given the number of CSVD-related comparisons, p-values from these models were additionally adjusted using the Benjamini–Hochberg false discovery rate procedure. Raw and FDR-adjusted p-values are reported. All analyses were conducted in RStudio (version 4.4.0).

## Results

### Prevalence of ToM deficits in Post-Stroke Aphasia

Table 3 summarizes ToM performance across the RUK and RK tasks. Classification was based on task-specific decision rules incorporating performance across false belief, true belief, and control trials. Performance was heterogeneous across both tasks. Most participants were classified as showing spared performance, but impaired performance was observed in a substantial subset of the sample. Impaired performance was more common on the RK task than on the RUK task, consistent with the greater inhibitory demands of the RK condition. Overall, these findings indicate that successful performance on nonverbal false-belief tasks is not uniformly preserved in post-stroke aphasia, even when linguistic demands are minimized. Individual-level task performance is reported in Supplementary Table 2.

**Table 3.** Summary of performance classifications on the two nonverbal false-belief tasks.

| ToM Performance Category | Reality-Unknown | Reality-Known |
| --- | --- | --- |
| Spared | 28 (64%) | 26 (59%) |
| Impaired | 10 (23%) | 16 (36%) |
| Processing difficulties | 3 (7%) | 2 (5%) |
| False negatives | 3 (7%) | — |
| Total N | 44 | 44 |

Classification was based on task-specific decision trees incorporating performance across false belief, true belief, and control trials, as described by Biervoye et al. (2018).

### Relationship of ToM deficits with Aphasia Severity

To examine whether aphasia severity predicts ToM performance, we ran linear regression models with WAB-R AQ scores as the predictor and percent accuracy on each ToM task as the outcome. As shown on Table 4, aphasia severity was not significantly associated with ToM accuracy for either the RUK (p = 0.34) or RK task (p = 0.58). These results remained unchanged when adjusting for age and lesion volume. Visualizations of these relationships are shown in Figure 1. Together, these findings suggest that aphasia severity, as measured by WAB-R AQ, is not a strong predictor of ToM performance, supporting the view that ToM deficits may arise independently of linguistic ability.

**Table 4.** Associations of aphasia severity, lesion volume, and age with performance on the two nonverbal false-belief tasks.

| **Predictor** | **Reality Unknown** |  |  |  | **Reality Known** |  |  |
| --- | --- | --- | --- | --- | --- | --- | --- |
|  | **Accuracy Estimate (SE)** | **t** | **p** |  | **Accuracy Estimate (SE)** | **t** | **p** |
| Intercept | 0.85 (0.21) | 4.00 | <0.001 |  | 0.83 (0.31) | 2.68 | 0.01 |
| WAB-R AQ | 0.00 (0.00) | 0.97 | 0.34 |  | 0.00 (0.00) | 0.55 | 0.58 |
| Lesion volume | −0.00 (0.00) | −0.10 | 0.93 |  | 0.00 (0.00) | 0.21 | 0.84 |
| Age | −0.00 (0.00) | −1.33 | 0.19 |  | −0.00 (0.00) | −0.79 | 0.43 |

### Influence of CSVD on ToM Task Performance

Results from linear regression models examining associations between CSVD markers and ToM task accuracy (RUK and RK), adjusted for age and lesion volume, are summarized in Table 5 and Supplementary Tables 3–11. Reported p values are unadjusted; q values reflect Benjamini– Hochberg false discovery rate correction across all CSVD-related comparisons. Overall, most CSVD markers were not significantly associated with task accuracy after correction for multiple comparisons.

**Table 5.** Association between CSVD markers and ToM task accuracy.

| CSVD marker | RUK $\beta$<br>(SE) | RUK $p$ | RUK $q$ | RK $\beta$<br>(SE) | RK $p$ | RK $q$ | N |
| --- | --- | --- | --- | --- | --- | --- | --- |
| Periventricular WMH | -0.04<br>(0.04) | 0.30 | 0.48 | -0.01<br>(0.04) | 0.81 | 0.89 | 31 |
| Deep WMH | -0.02<br>(0.03) | 0.57 | 0.74 | -0.05<br>(0.04) | 0.19 | 0.37 | 31 |
| Basal Ganglia EPVS | -0.11<br>(0.04) | **0.01** | **0.03** | -0.11<br>(0.05) | **0.02** | 0.09 | 28 |
| Centrum Semiovale EPVS | -0.08<br>(0.04) | **0.07** | 0.18 | -0.00<br>(0.06) | 0.95 | 0.95 | 28 |
| Lacunes (presence) | -0.08<br>(0.06) | 0.21 | 0.37 | 0.02<br>(0.08) | 0.84 | 0.90 | 31 |
| Lacunes count | -0.09<br>(0.03) | **0.01** | 0.07 | -0.05<br>(0.05) | 0.33 | 0.49 | 31 |
| CMB (presence) | -0.02<br>(0.06) | 0.72 | 0.87 | -0.16<br>(0.08) | 0.05 | 0.15 | 32 |
| CMB count | -0.05<br>(0.01) | **0.002** | **0.02** | -0.07<br>(0.02) | **<0.001** | **0.02** | 32 |
| Total CSVD score | -0.04<br>(0.03) | 0.20 | 0.37 | -0.03<br>(0.04) | 0.42 | 0.58 | 28 |

WMH: white matter hyperintensities; EPVS: enlarged perivascular spaces; CMB: cerebral microbleeds; CSVD: cerebral small vessel disease; RUK: reality unknown; RK: reality known.

Neither PVWMH nor DWMH (Fazekas scores 0–3) were associated with accuracy on either the RUK or RK task. Similarly, total CSVD burden (0-4 composite score) was not significantly related to performance on either task. Lacunes, when modeled as a binary variable (presence vs absence), also showed no significant association with task accuracy. Lacune count was negatively associated with RUK accuracy at the unadjusted level (p = 0.01), but this association did not remain significant after FDR correction (q = 0.07). No association was observed between lacune count and RK accuracy.

Among the individual CSVD markers, EPVS-BG and CMB count showed the clearest relationships with task performance. Higher EPVS-BG scores were associated with lower RUK accuracy (p = 0.01, q = 0.03), and this association remained significant after FDR correction. A similar negative association was observed for RK accuracy at the unadjusted level (p = 0.02), but this did not survive correction (q = 0.09). No significant effects were observed for centrum semiovale EPVS. Higher CMB count was associated with lower accuracy on both tasks, and these associations remained significant after FDR correction (RUK: p = 0.002, q = 0.02; RK: p < 0.001, q = 0.02). In contrast, CMB presence as a binary variable was not significantly associated with RUK accuracy and showed only a nominal association with RK accuracy (p = 0.05), which did not remain significant after correction (q = 0.15).

Taken together, these findings suggest that among the CSVD markers examined, greater basal ganglia EPVS burden and especially higher CMB count were most consistently associated with poorer performance on nonverbal false-belief tasks, independent of age and lesion volume.

## Discussion

The purpose of this study was to examine performance on two nonverbal false-belief tasks in individuals with post-stroke aphasia, following left-hemisphere stroke, and ask whether variability in task performance was related to aphasia severity and vascular brain health. Three main findings emerged. First, performance was heterogeneous across both tasks, with reduced performance observed in a substantial subset of participants and occurring more often on the Reality-Known (RK) task than on the Reality-Unknown (RUK) task. Second, aphasia severity was not significantly associated with performance on either task. Third, among the CSVD markers examined, cerebral microbleed (CMB) count showed the most consistent association with lower task accuracy, while basal ganglia EPVS showed a more selective association. Together, these findings suggest that reduced performance on nonverbal ToM tasks in post-stroke aphasia may reflect multiple interacting influences, including task demands and vascular brain health, rather than being attributable to aphasia severity.

At a broad level, our results indicate that successful performance on nonverbal false-belief tasks is not uniformly preserved in post-stroke aphasia, even when overt linguistic demands are minimized. Similar to earlier case studies,^7, 8, 29^ several participants in our sample performed at or near ceiling on both tasks, consistent with the view that mental state reasoning can remain relatively preserved in at least some individuals with aphasia under low-language testing conditions. At the same time, the group-level data revealed substantial variability. On the RUK task, 23% of participants were classified as impaired, whereas on the RK task 36% were classified as impaired. This pattern suggests that moving beyond single-case reports is important for capturing the broader range of performance seen in aphasia and for identifying the factors that may contribute to this variability.

The higher frequency of reduced performance on the RK task is also informative. Compared with the RUK condition, the RK condition places greater demands on self-perspective inhibition, because participants must suppress their own knowledge of reality while reasoning about another person’s false belief. Poorer performance in this condition is therefore consistent with the idea that success on nonverbal false-belief tasks depends not only on mental-state attribution, but also on domain-general processes such as inhibition, attentional control, and cognitive flexibility. This point is central to the interpretation of the present findings. Even though these tasks are widely used to assess ToM, they are not process-pure measures of mental state reasoning^15, 30, 31^. Accordingly, reduced performance on such tasks may be clinically meaningful without mapping one-to-one onto a primary impairment in ToM itself.

Aphasia severity did not significantly predict performance on either task. This finding suggests that language impairment alone does not fully explain variability in false-belief performance in this population. Individual variability was evident: some participants with more severe aphasia exhibited spared performance, whereas others with milder aphasia performed poorly. This pattern is broadly consistent with prior work suggesting at least partial dissociation between language and mental state reasoning systems. Specifically, mental state reasoning is supported by a distributed network of regions that includes the bilateral temporo-parietal junction and a set of regions along the cortical midline, areas consistently implicated in ToM across both verbal and nonverbal tasks.^32–34^ These regions are thought to represent others’ beliefs and intentions in a modality-independent format and are functionally distinct from the left-lateralized frontotemporal network that supports core higher-order language processing.^35, 36^ Recent fMRI evidence further supports this dissociation: Shain et al.^37^ demonstrated that the language-selective network shows minimal engagement during false-belief reasoning, even when tasks involve rich narrative content. Our findings reinforce the view that ToM and language rely on distinct neural systems: linguistic impairment does not necessarily imply deficits in social-cognitive functioning, and conversely, preserved language abilities do not guarantee intact ToM. Our findings also highlight the need for direct ToM assessment in PWA, rather than inferring social cognitive abilities from language severity alone.

The most novel aspect of the study concerns the role of vascular brain health. Among the CSVD markers examined, CMB count showed the most robust and consistent association with task performance, with higher CMB burden associated with lower accuracy on both RUK and RK even after correction for multiple comparisons. Basal ganglia EPVS also showed a significant association with lower RUK accuracy, although the corresponding association with RK accuracy did not survive correction. One plausible interpretation is that these CSVD markers index disruption to distributed neural systems that support executive control, information integration, and efficient coordination across brain regions, all of which are likely important for successful false-belief reasoning. EPVS are a common neuroimaging marker of CSVD and have been linked to executive dysfunction and global cognitive decline.^38^ Our findings extend this work by demonstrating that greater EPVS burden in the basal ganglia was associated with reduced accuracy on both ToM tasks, suggesting that EPVS-related disruption may impact executive processes such as inhibition and mental flexibility that are critical for belief reasoning.^39, 40^ Similarly, greater CMB burden was associated with poorer ToM accuracy in both tasks, reinforcing prior evidence that CMB are related cognitive impairment beyond memory deficits.^41, 42^ Finally, although lacunes were present in only a subset of participants, we observed a negative association between lacune count and ToM accuracy in the RUK task, suggesting that even a relatively small burden of lacunar infarcts, typically associated with impairments in executive function, processing speed, and attention,^43, 44^ may affect belief reasoning in some individuals. No significant associations were observed for WMH or total CSVD burden. This may reflect limited WMH severity in our relatively young sample (mean age: 57; only few participants had severe WMH, i.e., a Fazekas score of 3), consistent with evidence that WMH-related cognitive effects tend to emerge later in life^45^and are more pronounced in individuals with extensive lesion load.^26^ Moreover, total CSVD burden scores may mask the specific contributions of individual markers, especially when those markers differentially affect distinct cognitive domains and/or when the burden of pathology is mild to moderate.^46^

Taken together, these findings suggest that diffuse vascular pathology beyond the index stroke may contribute to variability in performance on socially demanding cognitive tasks in aphasia. One plausible interpretation is that these CSVD markers index disruption to distributed neural systems that support executive control, information integration, and efficient coordination across brain regions, all of which are likely important for successful false-belief reasoning. In this context, poorer performance on false-belief tasks may reflect the downstream behavioral consequences of vascular brain injury affecting the broader cognitive architecture required to perform the task, rather than selective disruption of mental-state reasoning per se. This interpretation fits well with the stronger impairment observed on the more inhibitory demanding RK task and helps explain why vascular burden may be particularly relevant even in paradigms designed to minimize language demands.

These findings have several clinical implications. First, they suggest that reduced performance on tasks intended to assess ToM in aphasia should be interpreted cautiously. Such difficulties may reflect not only social-cognitive processes, but also broader executive and vascular influences on task performance. Second, they underscore the importance of evaluating social-cognitive functioning directly rather than inferring it from aphasia severity alone. Finally, they raise the possibility that vascular comorbidity contributes to subtle but functionally meaningful communication difficulties that are not captured by standard aphasia measures. This may be particularly relevant in older stroke survivors, among whom CSVD is common.

Several limitations should be acknowledged. First, although the tasks minimized overt language demands, they do not isolate ToM independently of executive processes. As a result, the present study cannot determine whether reduced performance reflects impairment in mental state reasoning itself, in supporting domain-general mechanisms, or both. Second, while we used age-matched norms from Biervoye et al.^14^ to contextualize participant performance, we did not include a direct control group. Given that these norms were collected under similar conditions, we believe they provide a reasonable benchmark, though future studies would benefit from direct within-study comparisons to ensure full methodological consistency. Third, MRI data were not available for all participants who completed the behavioral tasks, and not all CSVD markers could be rated in every participant because different MRI sequences were required. This reduced sample size for some analyses and affected the derivation of total CSVD burden scores. Fourth, some CSVD findings were modest and did not survive correction for multiple comparisons. Finally, the cross-sectional design does not allow conclusions about causality or about the temporal relationship between vascular burden and false-belief task performance.

In summary, performance on nonverbal false-belief tasks in post-stroke aphasia was heterogeneous and was not fully explained by aphasia severity alone. Instead, selected markers of CSVD, particularly CMB count, were associated with lower task accuracy, supporting the view that broader vascular brain health contributes to variability in social-cognitive task performance after stroke. These findings argue for a more cautious interpretation of apparent ToM-related difficulties in aphasia and suggest that reduced performance on such tasks may reflect the combined effects of language impairment, executive demands, and vascular brain injury rather than a primary deficit in mental state reasoning and/or aphasia alone.

## Data Availability

All data produced in the present study are available upon reasonable request to the authors

## Acknowledgements

The authors would like to extend their gratitude to the research participants, for whom without this research would not be possible.

## Supplemental Material (List)

### MRI Acquisition Parameters & CSVD scoring

Multimodal MRI data were collected on 3T Siemens MAGNETOM Prisma scanners with 64-channel head coils at Boston University. The protocol included: a T1-weighted MEMPRAGE sequence (176 sagittal slices, 1.0 mm isotropic, TR = 2530 ms, TE1/TE2/TE3/TE4 = 1.69/3.55/5.41/7.27 ms, flip angle = 7°); a 3D T2-FLAIR SPACE sequence (176 sagittal slices, 1.0 mm isotropic, TR = 6000 ms, TE = 456 ms, flip angle = 7°); and a susceptibility-weighted imaging (SWI) GRE sequence (80 axial slices, 0.9 × 0.9 × 1.5 mm, TR = 27 ms, TE = 20 ms, flip angle = 15°, GRAPPA = 2).

CSVD markers will be rated according to Standards for Reporting Vascular Changes on Neuroimaging (STRIVE)^47^ guidelines and validated visual rating scales. All ratings will be performed blinded to demographic and clinical information. CSVD markers will include white matter hyperintensities, enlarged perivascular spaces, lacunes, and cerebral microbleeds. The severity of <u>white matter hyperintensities</u> in deep and periventricular regions will be evaluated on T2-FLAIR images using the Fazekas scale (range 0-3)^48^. <u>Enlarged perivascular spaces</u>, cerebrospinal fluid-filled cavities around small cerebral arterioles in the basal ganglia and centrum semiovale, will be rated on axial T2-FLAIR images using a validated 4-point scale. The presence and number of <u>lacunes</u> will be defined on T1-weighted images as hypointense lesions with a hyperintense rim^47^. <u>Cerebral microbleeds</u> presence and number will be evaluated on axial SWI images using current consensus criteria^49^. Following prior research^50^, the <u>total CSVD burden</u> will be determined on an ordinal scale from 0 to 4, assigning 1 point for each of the following: (1) moderate white matter hyperintensities (WMH) (periventricular WMH Fazekas 3 and/or deep WMH Fazekas 2-3); (2) moderate-to-severe enlarged perivascular spaces (EPVS) in the basal ganglia (>20); (3) presence of lacunes (≥1); and (4) presence of microbleeds (≥1).

**Supplementary Table 1.**
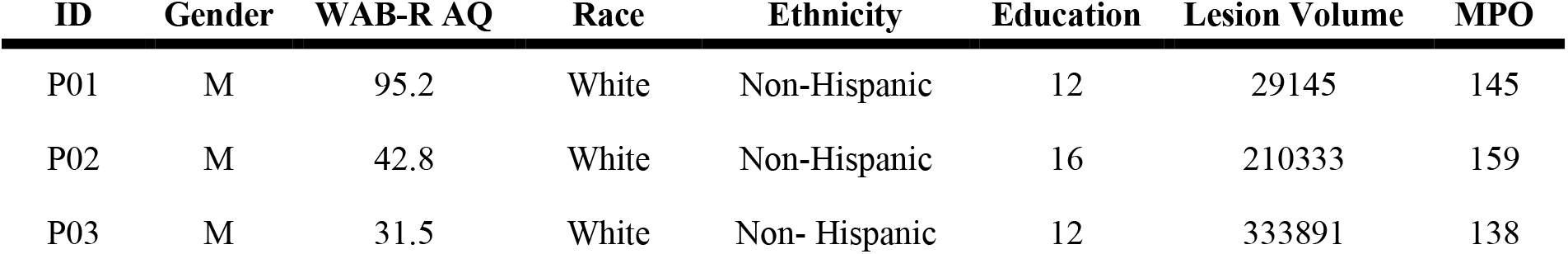
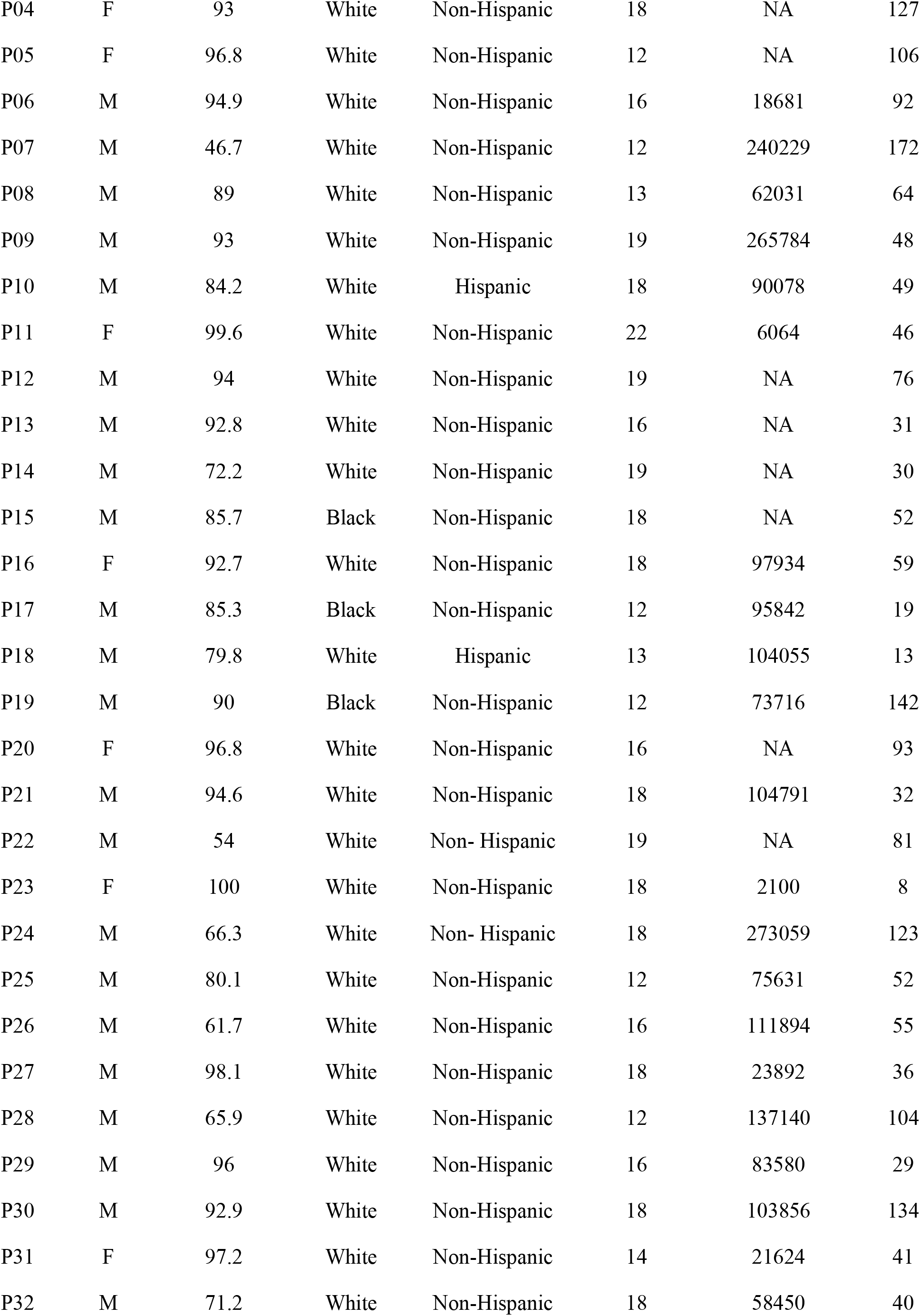
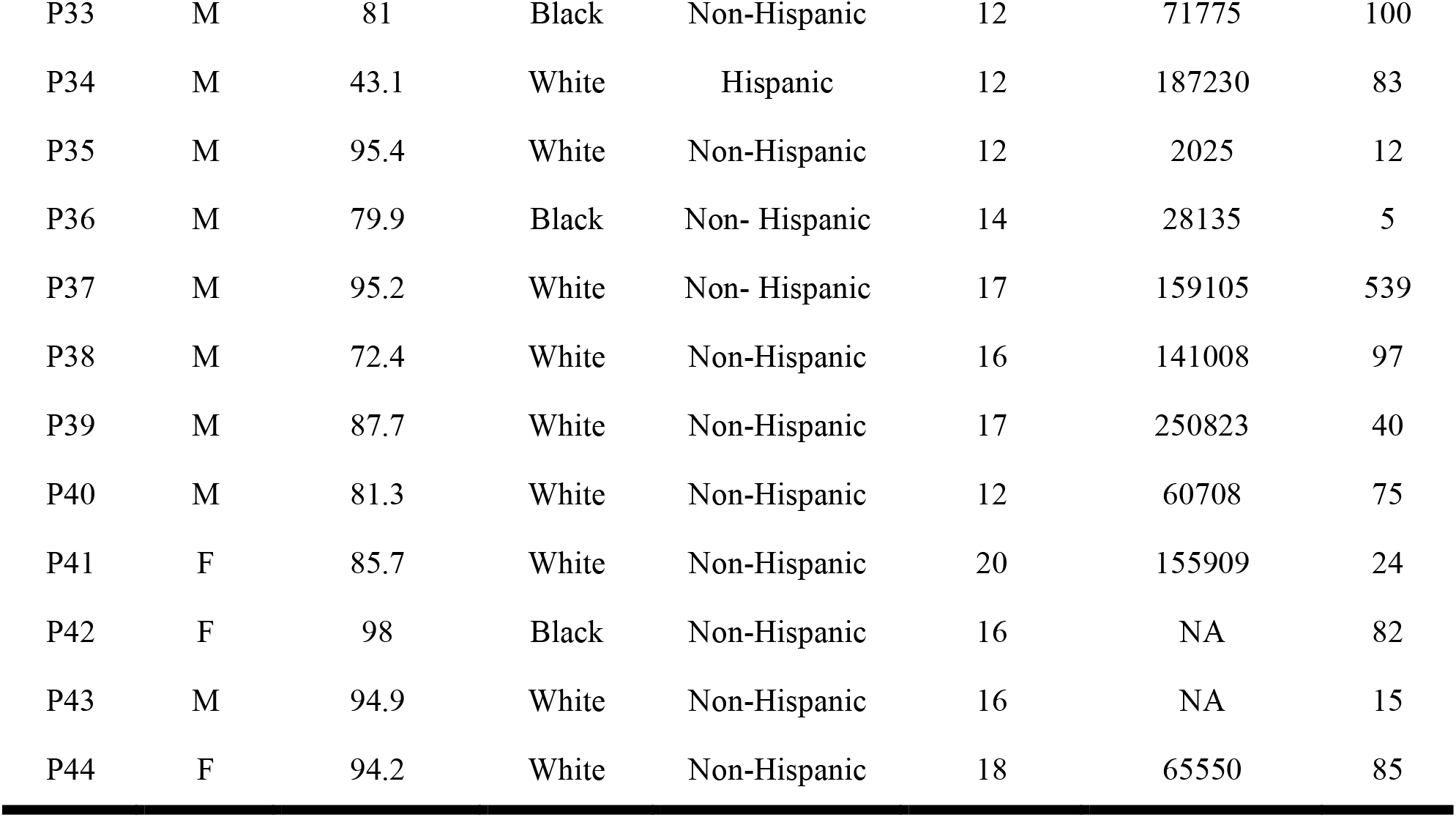
Individual-level participant characteristicss.

| ID | Gender | WAB-R AQ | Race | Ethnicity | Education | Lesion Volume | MPO |
| --- | --- | --- | --- | --- | --- | --- | --- |
| P01 | M | 95.2 | White | Non-Hispanic | 12 | 29145 | 145 |
| P02 | M | 42.8 | White | Non-Hispanic | 16 | 210333 | 159 |
| P03 | M | 31.5 | White | Non- Hispanic | 12 | 333891 | 138 |
| P04 | F | 93 | White | Non-Hispanic | 18 | NA | 127 |
| P05 | F | 96.8 | White | Non-Hispanic | 12 | NA | 106 |
| P06 | M | 94.9 | White | Non-Hispanic | 16 | 18681 | 92 |
| P07 | M | 46.7 | White | Non-Hispanic | 12 | 240229 | 172 |
| P08 | M | 89 | White | Non-Hispanic | 13 | 62031 | 64 |
| P09 | M | 93 | White | Non-Hispanic | 19 | 265784 | 48 |
| P10 | M | 84.2 | White | Hispanic | 18 | 90078 | 49 |
| P11 | F | 99.6 | White | Non-Hispanic | 22 | 6064 | 46 |
| P12 | M | 94 | White | Non-Hispanic | 19 | NA | 76 |
| P13 | M | 92.8 | White | Non-Hispanic | 16 | NA | 31 |
| P14 | M | 72.2 | White | Non-Hispanic | 19 | NA | 30 |
| P15 | M | 85.7 | Black | Non-Hispanic | 18 | NA | 52 |
| P16 | F | 92.7 | White | Non-Hispanic | 18 | 97934 | 59 |
| P17 | M | 85.3 | Black | Non-Hispanic | 12 | 95842 | 19 |
| P18 | M | 79.8 | White | Hispanic | 13 | 104055 | 13 |
| P19 | M | 90 | Black | Non-Hispanic | 12 | 73716 | 142 |
| P20 | F | 96.8 | White | Non-Hispanic | 16 | NA | 93 |
| P21 | M | 94.6 | White | Non-Hispanic | 18 | 104791 | 32 |
| P22 | M | 54 | White | Non- Hispanic | 19 | NA | 81 |
| P23 | F | 100 | White | Non-Hispanic | 18 | 2100 | 8 |
| P24 | M | 66.3 | White | Non- Hispanic | 18 | 273059 | 123 |
| P25 | M | 80.1 | White | Non-Hispanic | 12 | 75631 | 52 |
| P26 | M | 61.7 | White | Non-Hispanic | 16 | 111894 | 55 |
| P27 | M | 98.1 | White | Non-Hispanic | 18 | 23892 | 36 |
| P28 | M | 65.9 | White | Non-Hispanic | 12 | 137140 | 104 |
| P29 | M | 96 | White | Non-Hispanic | 16 | 83580 | 29 |
| P30 | M | 92.9 | White | Non-Hispanic | 18 | 103856 | 134 |
| P31 | F | 97.2 | White | Non-Hispanic | 14 | 21624 | 41 |
| P32 | M | 71.2 | White | Non-Hispanic | 18 | 58450 | 40 |
| P33 | M | 81 | Black | Non-Hispanic | 12 | 71775 | 100 |
| P34 | M | 43.1 | White | Hispanic | 12 | 187230 | 83 |
| P35 | M | 95.4 | White | Non-Hispanic | 12 | 2025 | 12 |
| P36 | M | 79.9 | Black | Non- Hispanic | 14 | 28135 | 5 |
| P37 | M | 95.2 | White | Non- Hispanic | 17 | 159105 | 539 |
| P38 | M | 72.4 | White | Non-Hispanic | 16 | 141008 | 97 |
| P39 | M | 87.7 | White | Non-Hispanic | 17 | 250823 | 40 |
| P40 | M | 81.3 | White | Non-Hispanic | 12 | 60708 | 75 |
| P41 | F | 85.7 | White | Non-Hispanic | 20 | 155909 | 24 |
| P42 | F | 98 | Black | Non-Hispanic | 16 | NA | 82 |
| P43 | M | 94.9 | White | Non-Hispanic | 16 | NA | 15 |
| P44 | F | 94.2 | White | Non-Hispanic | 18 | 65550 | 85 |

**Supplementary Table 2.** Raw trial-level scores and derived Theory of Mind (ToM) performance classifications for the Reality-Unknown (RUK) and Reality-Known (RK) nonverbal false-belief tasks. Classifications were determined using the decision-tree approach described by Biervoye et al. (2018).

| Participant | Reality Unknown |  |  |  | Reality Known |  |  |  |
| --- | --- | --- | --- | --- | --- | --- | --- | --- |
|  | FB/8 | TB/4 | MC/F/12 | Classification | FB/8 | TB/9 | MC/F/16 | Classification |
| P01 | 7 | 4 | 11 | Spared | 7 | 9 | 16 | Spared |
| P02 | 5 | 2 | 7 | False negative | 4 | 7 | 11 | Impaired |
| P03 | 2 | 4 | 10 | Impaired | 7 | 9 | 16 | Spared |
| P04 | 5 | 4 | 12 | Spared | 6 | 9 | 16 | Spared |
| P05 | 8 | 4 | 12 | Spared | 8 | 9 | 16 | Spared |
| P06 | 5 | 3 | 11 | Spared | 8 | 7 | 14 | Spared |
| P07 | 4 | 3 | 10 | Spared | 3 | 9 | 11 | Impaired |
| P08 | 5 | 4 | 11 | Spared | 0 | 9 | 16 | Impaired |
| P09 | 1 | 2 | 12 | Processing difficulties | 1 | 9 | 16 | Impaired |
| P10 | 7 | 1 | 5 | False negative | 2 | 2 | 4 | Processing difficulties |
| P11 | 6 | 3 | 12 | Spared | 8 | 9 | 16 | Spared |
| P12 | 8 | 4 | 12 | Spared | 8 | 9 | 15 | Spared |
| P13 | 1 | 4 | 9 | Impaired | 2 | 9 | 16 | Impaired |
| P14 | 7 | 4 | 12 | Spared | 4 | 9 | 16 | Impaired |
| P15 | 0 | 4 | 10 | Impaired | 6 | 9 | 16 | Spared |
| P16 | 8 | 4 | 12 | Spared | 8 | 8 | 15 | Spared |
| P17 | 6 | 4 | 10 | Spared | 7 | 9 | 16 | Spared |
| P18 | 6 | 4 | 12 | Spared | 7 | 9 | 14 | Spared |
| P19 | 6 | 4 | 9 | Spared | 3 | 9 | 15 | Impaired |
| P20 | 8 | 4 | 12 | Spared | 8 | 9 | 16 | Spared |
| P21 | 7 | 4 | 11 | Spared | 8 | 9 | 16 | Spared |
| P22 | 7 | 2 | 10 | Spared | 3 | 9 | 15 | Impaired |
| P23 | 8 | 4 | 12 | Spared | 8 | 9 | 16 | Spared |
| P24 | 5 | 4 | 12 | Spared | 7 | 9 | 14 | Spared |
| P25 | 2 | 4 | 12 | Processing difficulties | 7 | 9 | 14 | Spared |
| P26 | 4 | 4 | 12 | Spared | 7 | 9 | 16 | Spared |
| P27 | 4 | 4 | 12 | Impaired | 6 | 9 | 15 | Spared |
| P28 | 8 | 4 | 12 | Spared | 6 | 9 | 15 | Spared |
| P29 | 8 | 3 | 11 | Spared | 8 | 9 | 16 | Spared |
| P30 | 8 | 3 | 10 | Spared | 0 | 0 | 1 | Processing difficulties |
| P31 | 2 | 4 | 11 | Impaired | 0 | 9 | 16 | Impaired |
| P32 | 8 | 4 | 12 | Spared | 7 | 9 | 16 | Spared |
| P33 | 5 | 4 | 11 | Spared | 6 | 9 | 16 | Spared |
| P34 | 7 | 4 | 12 | Spared | 0 | 9 | 16 | Impaired |
| P35 | 8 | 4 | 12 | Spared | 8 | 9 | 16 | Spared |
| P36 | 3 | 3 | 7 | Processing difficulties | 0 | 9 | 16 | Impaired |
| P37 | 7 | 4 | 12 | Spared | 8 | 9 | 16 | Spared |
| P38 | 3 | 4 | 11 | Impaired | 6 | 9 | 15 | Spared |
| P39 | 4 | 2 | 12 | Impaired | 5 | 9 | 16 | Impaired |
| P40 | 0 | 4 | 2 | Impaired | 0 | 9 | 16 | Impaired |
| P41 | 7 | 4 | 12 | Spared | 7 | 9 | 15 | Spared |
| P42 | 0 | 4 | 12 | Impaired | 5 | 9 | 16 | Impaired |
| P43 | 1 | 2 | 10 | Impaired | 1 | 9 | 16 | Impaired |
| P44 | 7 | 1 | 11 | False negative | 1 | 8 | 15 | Impaired |

FB: false belief; TB: true belief; MC/F: memory control / filler

**Supplementary Table 3.**
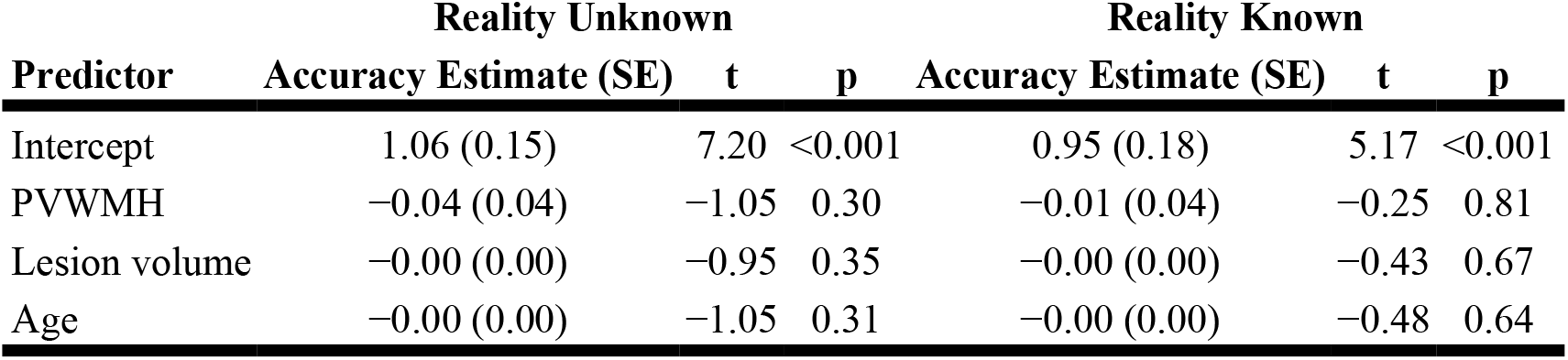
Association between Periventricular White Matter Hyperintensities (PVWMH) and ToM Accuracy.

| Predictor | Reality Unknown |  |  | Reality Known |  |  |
| --- | --- | --- | --- | --- | --- | --- |
|  | Accuracy Estimate (SE) | t | p | Accuracy Estimate (SE) | t | p |
| Intercept | 1.06 (0.15) | 7.20 | <0.001 | 0.95 (0.18) | 5.17 | <0.001 |
| PVWMH | -0.04 (0.04) | -1.05 | 0.30 | -0.01 (0.04) | -0.25 | 0.81 |
| Lesion volume | -0.00 (0.00) | -0.95 | 0.35 | -0.00 (0.00) | -0.43 | 0.67 |
| Age | -0.00 (0.00) | -1.05 | 0.31 | -0.00 (0.00) | -0.48 | 0.64 |

**Supplementary Table 4.**
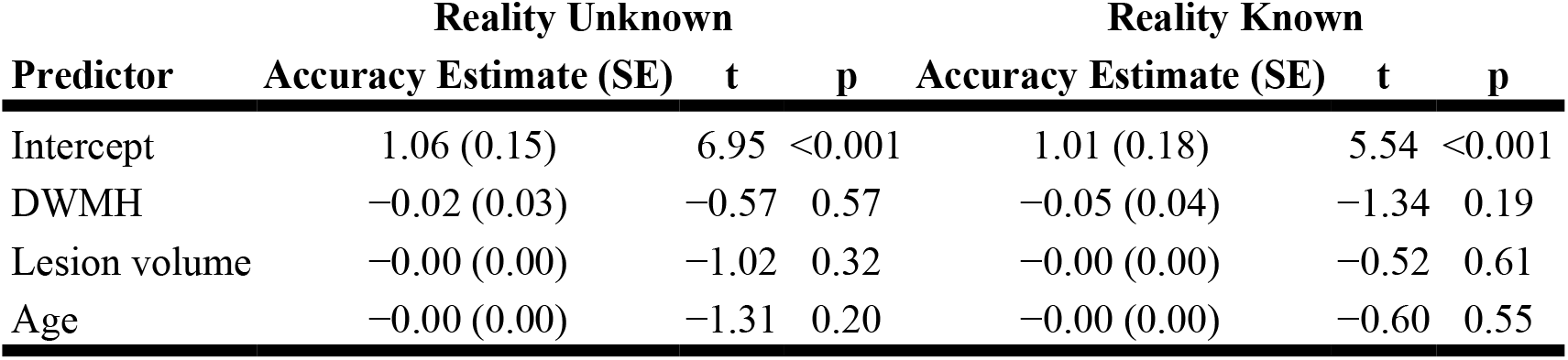
Association between Deep White Matter Hyperintensities (DWMH) and ToM Accuracy.

**Supplementary Table 5.** Association between Enlarged Perivascular Spaces in the Basal Ganglia (EPVS-BG) and ToM Accuracy.

| Predictor | Reality Unknown |  |  | Reality Known |  |  |
| --- | --- | --- | --- | --- | --- | --- |
|  | Accuracy Estimate (SE) | t | p | Accuracy Estimate (SE) | t | p |
| Intercept | 1.05 (0.13) | 8.03 | <0.001 | 0.99 (0.17) | 5.96 | <0.001 |
| EPVS-BG | -0.11 (0.04) | -3.06 | 0.01 | -0.11 (0.05) | -2.42 | 0.02 |
| Lesion volume | -0.00 (0.00) | -1.60 | 0.12 | -0.00 (0.00) | -0.67 | 0.51 |
| Age | -0.00 (0.00) | -0.07 | 0.94 | 0.00 (0.00) | 0.19 | 0.85 |

**Supplementary Table 6.** Association between Enlarged Perivascular Spaces in Centrum Semiovale (EPVS-CS) and ToM Accuracy.

| Predictor | Reality Unknown |  |  | Reality Known |  |  |
| --- | --- | --- | --- | --- | --- | --- |
|  | Accuracy Estimate (SE) | t | p | Accuracy Estimate (SE) | t | p |
| Intercept | 1.14 (0.16) | 7.37 | <0.001 | 0.98 (0.20) | 4.92 | <0.001 |
| EPVS-CS | -0.08 (0.04) | -1.87 | 0.07 | -0.00 (0.06) | -0.07 | 0.95 |
| Lesion volume | -0.00 (0.00) | -1.65 | 0.11 | -0.00 (0.00) | -0.35 | 0.73 |
| Age | -0.00 (0.00) | -0.63 | 0.54 | -0.00 (0.00) | -0.68 | 0.51 |

**Supplementary Table 7.**
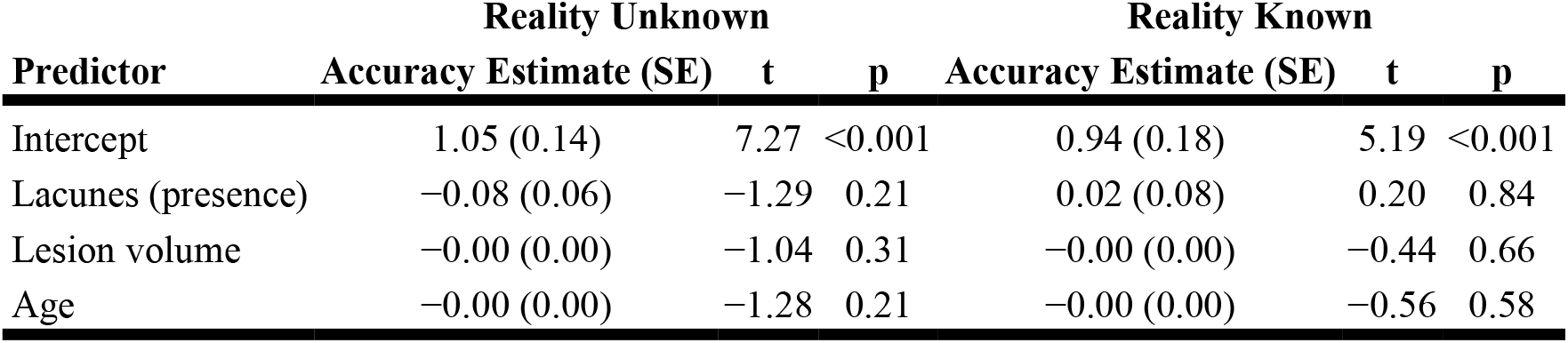
Association between Lacunes presence and ToM Accuracy.

| Predictor | Reality Unknown |  |  | Reality Known |  |  |
| --- | --- | --- | --- | --- | --- | --- |
|  | Accuracy Estimate (SE) | t | p | Accuracy Estimate (SE) | t | p |
| Intercept | 1.05 (0.14) | 7.27 | <0.001 | 0.94 (0.18) | 5.19 | <0.001 |
| Lacunes (presence) | -0.08 (0.06) | -1.29 | 0.21 | 0.02 (0.08) | 0.20 | 0.84 |
| Lesion volume | -0.00 (0.00) | -1.04 | 0.31 | -0.00 (0.00) | -0.44 | 0.66 |
| Age | -0.00 (0.00) | -1.28 | 0.21 | -0.00 (0.00) | -0.56 | 0.58 |

**Supplementary Table 8.** Association between Lacunes presence and ToM Accuracy.

| Predictor | Reality Unknown |  |  | Reality Known |  |  |
| --- | --- | --- | --- | --- | --- | --- |
|  | Accuracy Estimate (SE) | t | p | Accuracy Estimate (SE) | t | p |
| Intercept | 1.07 (0.13) | 8.03 | <0.001 | 0.96 (0.18) | 5.37 | <0.001 |
| Lacunes (count) | -0.09 (0.03) | -2.62 | 0.01 | -0.05 (0.05) | -1.00 | 0.33 |
| Lesion volume | -0.00 (0.00) | -1.50 | 0.15 | -0.00 (0.00) | -0.60 | 0.55 |
| Age | -0.00 (0.00) | -1.38 | 0.18 | -0.00 (0.00) | -0.53 | 0.60 |

**Supplementary Table 9.**
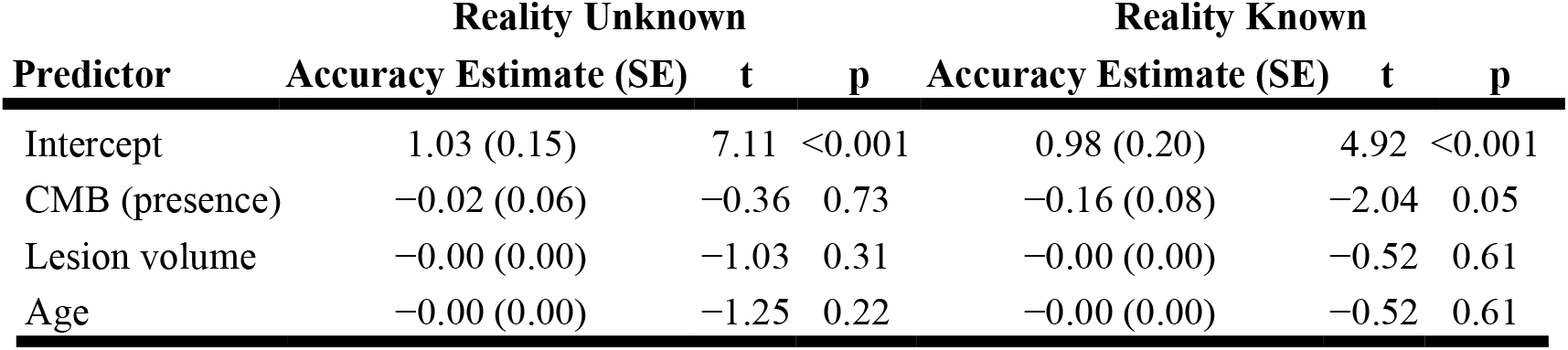
Association between Cerebral Microbleeds (CMB) presence and ToM Accuracy.

| Predictor | Reality Unknown |  |  | Reality Known |  |  |
| --- | --- | --- | --- | --- | --- | --- |
|  | Accuracy Estimate (SE) | t | p | Accuracy Estimate (SE) | t | p |

|  | Reality Unknown |  |  | Reality Known |  |  |
| --- | --- | --- | --- | --- | --- | --- |
| Intercept | 1.03 (0.15) | 7.11 | <0.001 | 0.98 (0.20) | 4.92 | <0.001 |
| CMB (presence) | -0.02 (0.06) | -0.36 | 0.73 | -0.16 (0.08) | -2.04 | 0.05 |
| Lesion volume | -0.00 (0.00) | -1.03 | 0.31 | -0.00 (0.00) | -0.52 | 0.61 |
| Age | -0.00 (0.00) | -1.25 | 0.22 | -0.00 (0.00) | -0.52 | 0.61 |

**Supplementary Table 10.** Association between Cerebral Microbleeds (CMB) count and ToM Accuracy.

|  | Reality Unknown |  |  | Reality Known |  |  |
| --- | --- | --- | --- | --- | --- | --- |
| Predictor | Accuracy Estimate (SE) | t | p | Accuracy Estimate (SE) | t | p |
| Intercept | 1.05 (0.12) | 8.52 | <0.001 | 0.98 (0.17) | 5.68 | <0.001 |
| CMB count | -0.05 (0.01) | -3.39 | 0.002 | -0.07 (0.02) | -3.79 | <0.001 |
| Lesion volume | -0.00 (0.00) | -1.91 | 0.07 | -0.00 (0.00) | -1.27 | 0.21 |
| Age | -0.00 (0.00) | -1.17 | 0.25 | -0.00 (0.00) | -0.41 | 0.69 |

**Supplementary Table 11.** Association between total Cerebral Small Vessel Disease (CSVD) burden and ToM Accuracy.

|  | Reality Unknown |  |  | Reality Known |  |  |
| --- | --- | --- | --- | --- | --- | --- |
| Predictor | Accuracy Estimate (SE) | t | p | Accuracy Estimate (SE) | t | p |
| Intercept | 1.05 (0.15) | 7.00 | <0.001 | 0.98 (0.18) | 5.37 | <0.001 |
| Total CSVD score | -0.04 (0.03) | -1.33 | 0.20 | -0.03 (0.04) | -0.83 | 0.42 |
| Lesion volume | -0.00 (0.00) | -1.29 | 0.21 | -0.00 (0.00) | -0.48 | 0.64 |
| Age | -0.00 (0.00) | -0.97 | 0.34 | -0.00 (0.00) | -0.60 | 0.56 |

## References

1. Weed E. Theory of mind impairment in right hemisphere damage: A review of the evidence. Int J Speech Lang Pathol 2008;10:414–424.

2. Tompkins CA, Scharp VL, Fassbinder W, Meigh KM, Armstrong EM. A different story on “Theory of Mind” deficit in adults with right hemisphere brain damage. Aphasiology 2008;22:42–61.

3. Balaban N, Friedmann N, Ziv M. Theory of mind impairment after right-hemisphere damage. Aphasiology 2016;30:1399–1423.

4. Schurz M, Radua J, Aichhorn M, Richlan F, Perner J. Fractionating theory of mind: A meta-analysis of functional brain imaging studies. Neuroscience & Biobehavioral Reviews 2014;42:9–34.

5. Wellman HM, Cross D, Watson J. Meta-analysis of theory-of-mind development: the truth about false belief. Child Dev 2001;72:655–684.

6. Apperly IA. What is “theory of mind”? Concepts, cognitive processes and individual differences. Q J Exp Psychol (Hove) 2012;65:825–839.

7. Varley R, Siegal M. Evidence for cognition without grammar from causal reasoning and ‘theory of mind’ in an agrammatic aphasic patient. Current Biology 2000;10:723–726.

8. Varley R, Siegal M, Want SC. Severe Impairment in Grammar Does Not Preclude Theory of Mind. Neurocase 2001;7:489–493.

9. Kiran S, Thompson CK. Neuroplasticity of Language Networks in Aphasia: Advances, Updates and Future Challenges. Frontiers in Neurology 2019;10:295–295.

10. Henry JD, Phillips LH, Ruffman T, Bailey PE. A meta-analytic review of age differences in theory of mind. Psychol Aging 2013;28:826–839.

11. Simmons-Mackie NN, Damico JS. Access and social inclusion in aphasia: Interactional principles and applications. Aphasiology 2007;21:81–97.

12. Cotter J, Granger K, Backx R, Hobbs M, Looi CY, Barnett JH. Social cognitive dysfunction as a clinical marker: A systematic review of meta-analyses across 30 clinical conditions. Neurosci Biobehav Rev 2018;84:92–99.

13. Davidson B, Howe T, Worrall L, Hickson L, Togher L. Social participation for older people with aphasia: the impact of communication disability on friendships. Top Stroke Rehabil 2008;15:325–340.

14. Biervoye A, Meert G, Apperly IA, Samson D. Assessing the integrity of the cognitive processes involved in belief reasoning by means of two nonverbal tasks: Rationale, normative data collection and illustration with brain-damaged patients. PLoS ONE 2018;13:1–29.

15. Aboulafia-Brakha T, Christe B, Martory MD, Annoni JM. Theory of mind tasks and executive functions: a systematic review of group studies in neurology. J Neuropsychol 2011;5:39–55.

16. Apperly IA, Samson D, Humphreys GW. Studies of adults can inform accounts of theory of mind development. Dev Psychol 2009;45:190–201.

17. Wade M, Prime H, Jenkins JM, Yeates KO, Williams T, Lee K. On the relation between theory of mind and executive functioning: A developmental cognitive neuroscience perspective. Psychon Bull Rev 2018;25:2119–2140.

18. Organization WH. 2023 [online]. Available at: https://www.who.int/publications/i/item/9789240054561.

19. German TP, Hehman JA. Representational and executive selection resources in ‘theory of mind’: Evidence from compromised belief-desire reasoning in old age. Cognition 2006;101:129–152.

20. Phillips LH, Bull R, Allen R, Insch P, Burr K, Ogg W. Lifespan aging and belief reasoning: Influences of executive function and social cue decoding. Cognition 2011;120:236–247.

21. Zhou W, Hong Z, Chen D, Liu S, Zhang L. The mechanism of inhibitory control on the development of theory of mind in old age—based on the two-component model of psychological theory. Aging & Mental Health 2021;25:341–349.

22. Duval C, Piolino P, Bejanin A, Eustache F, Desgranges B. Age effects on different components of theory of mind. Conscious Cogn 2011;20:627–642.

23. Wardlaw JM, Smith C, Dichgans M. Mechanisms of sporadic cerebral small vessel disease: Insights from neuroimaging. The Lancet Neurology 2013;12:483–497.

24. Bos D, Wolters FJ, Darweesh SKL, et al. Cerebral small vessel disease and the risk of dementia: A systematic review and meta-analysis of population-based evidence. Elsevier Inc., 2018: 1482–1492.

25. Huijts M, Duits A, Van Oostenbrugge RJ, Kroon AA, De Leeuw PW, Staals J. Accumulation of MRI markers of cerebral small vessel disease is associated with decreased cognitive function. A study in first-ever lacunar stroke and hypertensive patients. Frontiers in Aging Neuroscience 2013;5:1–7.

26. Kynast J, Lampe L, Luck T, et al. White matter hyperintensities associated with small vessel disease impair social cognition beside attention and memory. Journal of Cerebral Blood Flow & Metabolism 2018;38:996–1009.

27. Kertesz A. Western aphasia battery (Revised). San Antonio, TX: Psychological Corp., 2007.

28. Wardlaw JM, Smith EE, Biessels GJ, et al. Neuroimaging standards for research into small vessel disease and its contribution to ageing and neurodegeneration. The Lancet Neurology 2013;12:822–838.

29. Apperly IA, Samson D, Carroll N, Hussain S, Humphreys G. Intact first- and second-order false belief reasoning in a patient with severely impaired grammar. Soc Neurosci 2006;1:334–348.

30. Quesque F, Rossetti Y. What Do Theory-of-Mind Tasks Actually Measure? Theory and Practice. Perspect Psychol Sci 2020;15:384–396.

31. Siegal M, Varley R. Aphasia, language, and theory of mind. Social neuroscience 2006;1:167–174.

32. Saxe R, Kanwisher N. People thinking about thinking people. The role of the temporo-parietal junction in “theory of mind”. Neuroimage 2003;19:1835–1842.

33. Jacoby N, Bruneau E, Koster-Hale J, Saxe R. Localizing Pain Matrix and Theory of Mind networks with both verbal and non-verbal stimuli. Neuroimage 2016;126:39–48.

34. Gallagher HL, Happé F, Brunswick N, Fletcher PC, Frith U, Frith CD. Reading the mind in cartoons and stories: an fMRI study of ‘theory of mind’ in verbal and nonverbal tasks. Neuropsychologia 2000;38:11–21.

35. Binder JR, Frost JA, Hammeke TA, Cox RW, Rao SM, Prieto T. Human brain language areas identified by functional magnetic resonance imaging. J Neurosci 1997;17:353–362.

36. Fedorenko E, Hsieh P-J, Nieto-Castañón A, Whitfield-Gabrieli S, Kanwisher N. New Method for fMRI Investigations of Language: Defining ROIs Functionally in Individual Subjects. Journal of Neurophysiology 2010;104:1177–1194.

37. Shain C, Paunov A, Chen X, Lipkin B, Fedorenko E. No evidence of theory of mind reasoning in the human language network. Cereb Cortex 2023;33:6299–6319.

38. Huijts M, Duits A, Staals J, Kroon AA, de Leeuw PW, van Oostenbrugge RJ. Basal ganglia enlarged perivascular spaces are linked to cognitive function in patients with cerebral small vessel disease. Curr Neurovasc Res 2014;11:136–141.

39. Hamilton J, Radlak B, Morris PG, Phillips LH. Theory of Mind and Executive Functioning Following Stroke. Arch Clin Neuropsychol 2017;32:507–518.

40. Pluta A, Gawron N, Sobańska M, Wójcik AD, Łojek E. The nature of the relationship between neurocognition and theory of mind impairments in stroke patients. Neuropsychology 2017;31:666–681.

41. van Norden AG, van den Berg HA, de Laat KF, Gons RA, van Dijk EJ, de Leeuw FE. Frontal and temporal microbleeds are related to cognitive function: the Radboud University Nijmegen Diffusion Tensor and Magnetic Resonance Cohort (RUN DMC) Study. Stroke 2011;42:3382–3386.

42. Poels MMF, Ikram MA, van der Lugt A, et al. Cerebral microbleeds are associated with worse cognitive function. Neurology 2012;78:326–333.

43. Jokinen H, Kalska H, Ylikoski R, et al. Longitudinal cognitive decline in subcortical ischemic vascular disease - the ladis study. Cerebrovascular Diseases 2009;27:384–391.

44. Tuladhar AM, Reid AT, Shumskaya E, et al. Relationship between white matter hyperintensities, cortical thickness, and cognition. Stroke 2015;46:425–432.

45. Rodriguez L, Araujo AT, D DV, et al. Prevalence and imaging characteristics of cerebral small vessel disease in a Colombian population aged 40 years and older. Brain Commun 2024;6:fcae057.

46. Del Brutto VJ, Ortiz JG, Del Brutto OH, Mera RM, Zambrano M, Biller J. Total cerebral small vessel disease score and cognitive performance in community-dwelling older adults. Results from the Atahualpa Project. International Journal of Geriatric Psychiatry 2018;33:325–331.

47. Wardlaw JM, Smith EE, Biessels GJ, et al. Neuroimaging standards for research into small vessel disease and its contribution to ageing and neurodegeneration. Lancet Neurology 2013;12:822–838.

48. Fazekas F, Chawluk JB, Alavi A, Hurtig HI, Zimmerman RA. MR Signal Abnormalities at 1. 5 T in Alzheimer’s Dementia and Normal Aging deficiency. American Journal of Roentgenology 1987;149:351–356.

49. Greenberg SM, Vernooij MW, Cordonnier C, et al. Cerebral microbleeds: a guide to detection and interpretation. The Lancet Neurology 2009;8:165–174.

50. Pasi M, Sugita L, Xiong L, et al. Association of Cerebral Small Vessel Disease and Cognitive Decline After Intracerebral Hemorrhage 2021.

